# Fast Organ-of-Origin Classification for Digital Pathology Quality Control

**DOI:** 10.64898/2026.02.03.26345443

**Authors:** Witali Aswolinskiy, John K.L. Wong, Myroslav Zapukhlyak, Yulia Kindruk, Martin Paulikat, Christian Aichmüller

## Abstract

Digitizing large histopathology archives requires processing millions of scanned whole slide images that must be validated rapidly. Automated organ-of-origin classification can accelerate quality control and enable early detection of mislabeled specimens. We developed a deep learning model that classifies the organ of origin from H&E-stained slides using a single low-resolution thumbnail per slide in under one second. For training, we used thumbnails from 16,624 slides from the TCGA and CPTAC archives, which contain mostly primary tumor resections. The images were categorized into 14 classes based on the most common primary sites in TCGA: Bladder, Brain, Breast, Colorectal, Kidney, Liver, Lung, Pancreas, Prostate, Skin, Stomach, Thyroid gland, Uterus, and Other (encompassing the remaining tissue types). We evaluated our approach on two independent external cohorts: a 5-class cohort with 2,857 slides (Colorectal, Kidney, Liver, Pancreas, Prostate) and a comprehensive 14-class cohort (12,348 slides). The model achieved 90% balanced accuracy for the 5-class cohort and 62% for the full 14-class cohort. Notably, when considering only the predictions with high confidence, 53% of the large cohort could be classified with 74% balanced accuracy. Manual review of high-confidence misclassifications suggested that some may reflect errors in the ground truth rather than model error. Mean model inference time was 0.2s per slide on an NVIDIA L4 GPU. Our deep learning approach demonstrates high classification performance with very low inference time, indicating its potential for real-time and cost-effective quality control in digital pathology.

## 1 Introduction

Any hospital with a pathology archive will store tens of thousands of glass slides and paraffin blocks [1] and large cancer centers will even have millions [2]. Digitizing and analyzing this vast amount of data may yield breakthroughs in precision diagnostics that were not possible without the recent advances in large-scale computing. However, the required digitization brings practical challenges that hinder adoption of digital pathology. To take advantage of new AI tools, the large archives with millions of slides must not only be processed and stored, but also validated within strict operational timelines. One critical but often overlooked quality issue is specimen mislabeling, where slides become separated from their metadata or are scanned with incorrect identifiers. This can occur during scanning, file renaming or later data transfer between storage systems, and can lead to severe problems if the data is used for diagnostics without oversight [3, 4]. Early detection of such errors is essential to prevent misdiagnosis and ensure patient safety.

Our approach is to visually verify that the tissue visible on each H&E slide matches the expected organ of origin. However, this is a harder problem than it may appear. Slides may contain only limited organ-specific tissue (especially biopsies), include multiple tissue types, or be affected by artifacts. Some organs are histologically similar at low resolution, and in non-primary resections the sampled tissue may not match the primary site. For these reasons, we frame organ-of-origin prediction as a quality-control triage step rather than universal tissue identification.

The focus of automated quality control in histopathology has been on tissue quality: whether tissue is actually present on the slide and visible tissue artifacts, such as blur or ink [5, 6]. Prior work has also shown that low-resolution whole-slide thumbnails can already support fast triage tasks, for example normal vs malignant slide classification [7]. However, these approaches do not address organ-of-origin verification, which is a distinct quality-control problem: confirming that the scanned tissue matches the expected organ label in the metadata. Here, we focus on organ-of-origin classification from slide thumbnails to deploy as part of quality control for large digital pathology archives. Similar ideas have been explored in animal models, such as rat organ classification [8], but not systematically in human diagnostic workflows. Finding inconsistencies between expected and actual tissue origin before slides are further processed or integrated into clinical workflows can prevent costly re-scanning, reduce diagnostic turnaround time and detect contamination before it affects patient care.

In this work, we report a deep learning approach designed specifically for speed and practical deployment as an early triage step. We developed a model capable of classifying tissue origin from hematoxylin and eosin (H&E)-stained slides in under one second, enabling seamless integration into existing digital pathology workflows without processing delays. We demonstrate that such rapid, automated tissue classification can identify mislabeled or unexpected tissue types early in the pipeline, providing a cost-effective quality assurance mechanism. This bridges the gap between the recognized clinical need for specimen verification and the computational constraints of modern high-throughput pathology environments.

## 2. Materials and methods

### 2.1. Data

We collected digitized H&E-stained whole slide images (WSIs) from multiple public sources. Fourteen organ classes were defined based on the most common primary sites in The Cancer Genome Atlas (TCGA, [9]): Bladder, Brain, Breast, Colorectal, Kidney, Liver, Lung, Pancreas, Prostate, Skin, Stomach, Thyroid gland, Uterus (including Cervix), and *Other* (encompassing Adrenal gland, Esophagus, Heart, Larynx, Ovary, Soft Tissues, Peritoneum, Testis and Tongue). The model was originally trained using 23 organ classes; however, 9 classes with limited sample size and unstable performance were merged into the *Other* category, resulting in the 14-class setup used throughout this study. Full details of the sites and slide counts are provided in Supplementary Table S1.

For training, we used 11,579 diagnostic WSIs from TCGA and 5,045 slides from the Clinical Proteomic Tumor Analysis Consortium (CPTAC, [10]).

Evaluation was performed on two independent external datasets. The PAIP cohort [11] with 2,857 slides across five organ classes (Colorectal, Kidney, Liver, Pancreas, Prostate) and the VML cohort [12] with 12,348 slides representing all fourteen organ classes. Slide inclusion was based on the tissue area determined by contrast-based thresholding. Only slides with a largest tissue component of at least 1 mm^2^ were included.

Table 1 summarizes the number of whole slide images per organ across the different datasets used in this study.

**Table 1.** Number of whole slide images per organ and dataset.

| Organ | TCGA | CPTAC | PAIP | VML | Total |
| --- | --- | --- | --- | --- | --- |
| Bladder | 458 | 0 | 0 | 120 | 578 |
| Brain | 1706 | 244 | 0 | 427 | 2377 |
| Breast | 1122 | 396 | 0 | 1273 | 2791 |
| Colorectal | 602 | 223 | 893 | 529 | 2247 |
| Kidney | 944 | 777 | 400 | 546 | 2667 |
| Liver | 408 | 0 | 604 | 546 | 1558 |
| Lung | 1044 | 1526 | 0 | 583 | 3153 |
| Other | 2427 | 540 | 0 | 4905 | 7872 |
| Pancreas | 209 | 491 | 360 | 149 | 1209 |
| Prostate | 450 | 0 | 600 | 577 | 1627 |
| Skin | 418 | 0 | 0 | 821 | 1239 |
| Stomach | 379 | 0 | 0 | 178 | 557 |
| Thyroid gland | 508 | 0 | 0 | 478 | 986 |
| Uterus | 904 | 848 | 0 | 1216 | 2968 |
| Total | 11579 | 5045 | 2857 | 12348 | 31829 |

### 2.2. Training

For training, we extracted thumbnails from the WSIs at the coarsest pyramid level that yielded a minimum side length of 1024 pixels. This results in thumbnails taken at different magnifications and, effectively, multi-resolution training. The model architecture was based on ConvNeXt-Small [13] pretrained on ImageNet. We trained using a 5-fold cross-validation strategy with early stopping to prevent overfitting. As stopping metric, we used the Macro F1 score on the respective validation data. Data augmentation included random resized cropping, rotations, flips, brightness and contrast adjustments, blur operations, sharpening or embossing, and JPEG compression.

From the five trained models, two with the highest performance on the respective internal validation data were selected for evaluation to increase processing speed while improving generalization via ensembling.

### 2.3. Evaluation

For evaluation, rather than classifying the complete whole-slide thumbnail, we extracted the thumbnail corresponding to the largest tissue component within each slide. For this, we determined tissue using contrast-based thresholding in both HSV and Lab space and fitted a bounding box around the largest tissue component. This image is then extracted and rescaled so that its minimum side length is 1024. An overview of the approach is shown in Fig. 1.

**Figure 1.**
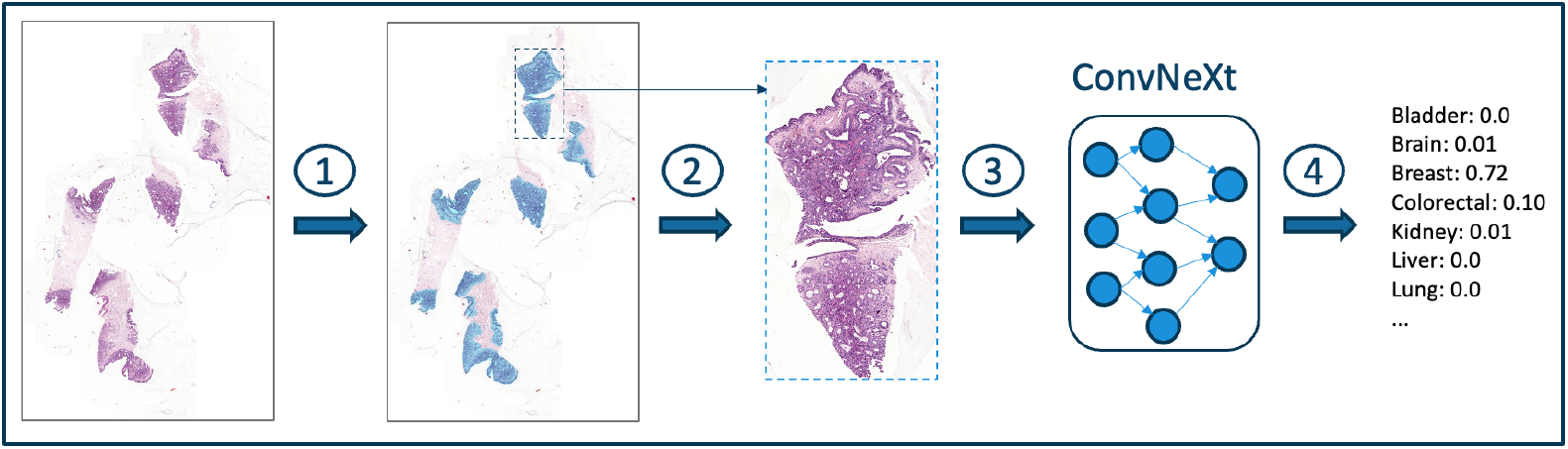
Method overview. 1: Tissue detection, 2: Extraction of thumbnail of largest tissue component, 3: Classification, 4: Maximum score is taken as prediction (“Breast” in the shown example).

## 3. Results

### 3.1. Speed evaluation

During inference, batch size was set to one, simulating sequential processing in a real workflow. On average, for the model forward pass (after thumbnail extraction) we measured 0.2s per image with an NVIDIA L4 GPU on a g6.xlarge AWS instance, and 1.4s on a CPU-only c6id.2xlarge AWS instance. Speed of thumbnail extraction relies predominantly on the storage medium (local vs cloud-based).

### 3.2. Performance evaluation

We evaluated our approach on the PAIP and VML cohorts. For VML, we first report overall performance on the full evaluation cohort (n = 12,348). In contrast to the training dataset, it also contains non-primary (metastatic) resections, biopsies, healthy tissue and combinations thereof. To increase clinical insight and interpretation, and to account for differences by specimen type, we also examine four subsets: primary tumor resections (n = 7,234), (ii) non-primary tumor resections (n = 2,965), (iii) healthy resections (n = 1,420) and (iv) tumor biopsies (n = 729). For each setting we compute the balanced accuracy and Macro F1 score, as these metrics reflect performance across all classes, not only those with the largest number of samples. The metrics along with 95% CIs (bootstrap with 1,000 resamples) are shown in Table 2. We also show the full confusion matrices to highlight organ-specific error patterns in Fig. 2.

**Table 2.** Classification performance (mean with 95% confidence interval).

| Dataset | Subset | N | Balanced Accuracy | Macro F1 |
| --- | --- | --- | --- | --- |
| PAIP | All | 2857 | 0.898 (0.887-0.909) | 0.928 (0.919-0.936) |
| VML | All | 12348 | 0.622 (0.609-0.634) | 0.564 (0.551-0.576) |
| VML | Primary Tumor Resections | 7234 | 0.665 (0.650-0.681) | 0.618 (0.603-0.634) |
| VML | Non-Primary Tumor Resections | 2965 | 0.651 (0.611-0.685) | 0.542 (0.511-0.567) |
| VML | Healthy Resections | 1420 | 0.508 (0.463-0.569) | 0.400 (0.369-0.435) |
| VML | Tumor Biopsies | 729 | 0.391 (0.305-0.454) | 0.272 (0.238-0.308) |

**Figure 2.**
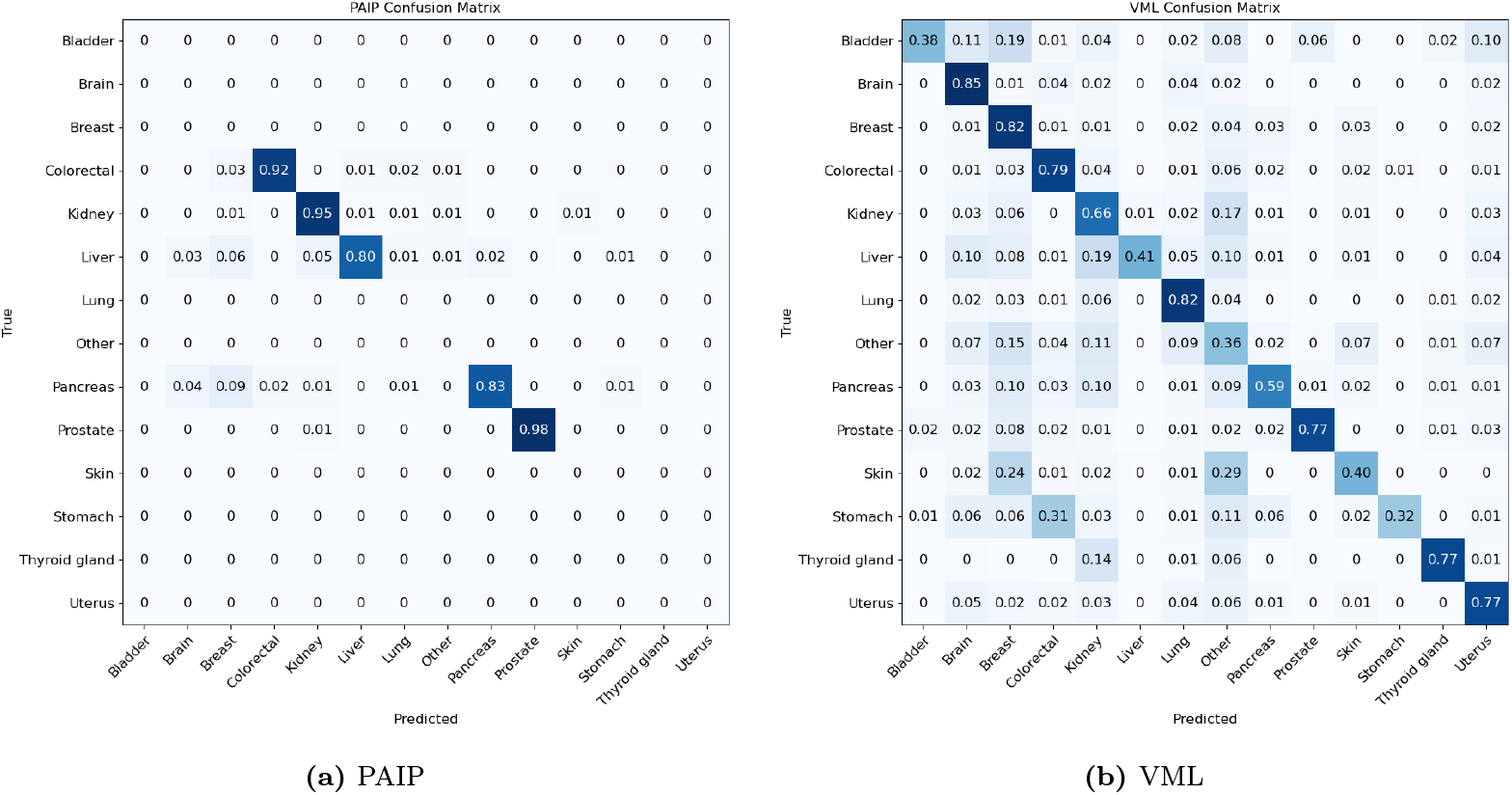
Normalized confusion matrices for PAIP (left) and VML (right) cohorts.

For PAIP (5 organs), the model achieved 90% balanced accuracy. For VML, the performance was lower, ranging from 39% (tumor biopsies) and 67% (primary tumor resections). Notably, performance on non-primary resections was only modestly lower than on primary resections. Healthy resections, however, and especially tumor biopsies, presented a larger challenge for the model.

#### 3.3. Probability cutoff evaluation

In multiclass classification, predictions are typically assigned to the class with the highest predicted probability. However, this leads to predictions with relatively low confidence. Therefore, we analyzed how applying a minimum probability cutoff affects model performance. Fig. 3 shows, for both evaluation cohorts, the impact on performance, when predictions below a given probability cutoff are discarded. With increasing cutoff, predictions become more reliable and balanced accuracy improves, at the cost of reduced coverage. For example, in the VML cohort, 74% balanced accuracy can be reached for approximately 53% of the samples at 0.8 probability cutoff. Notably, the performance improvement plateaus at higher cutoffs, suggesting an upper limit beyond which restricting predictions to the most confident samples does not yield further gains, potentially due to over-confident predictions or noise in the ground-truth labels.

**Figure 3.**
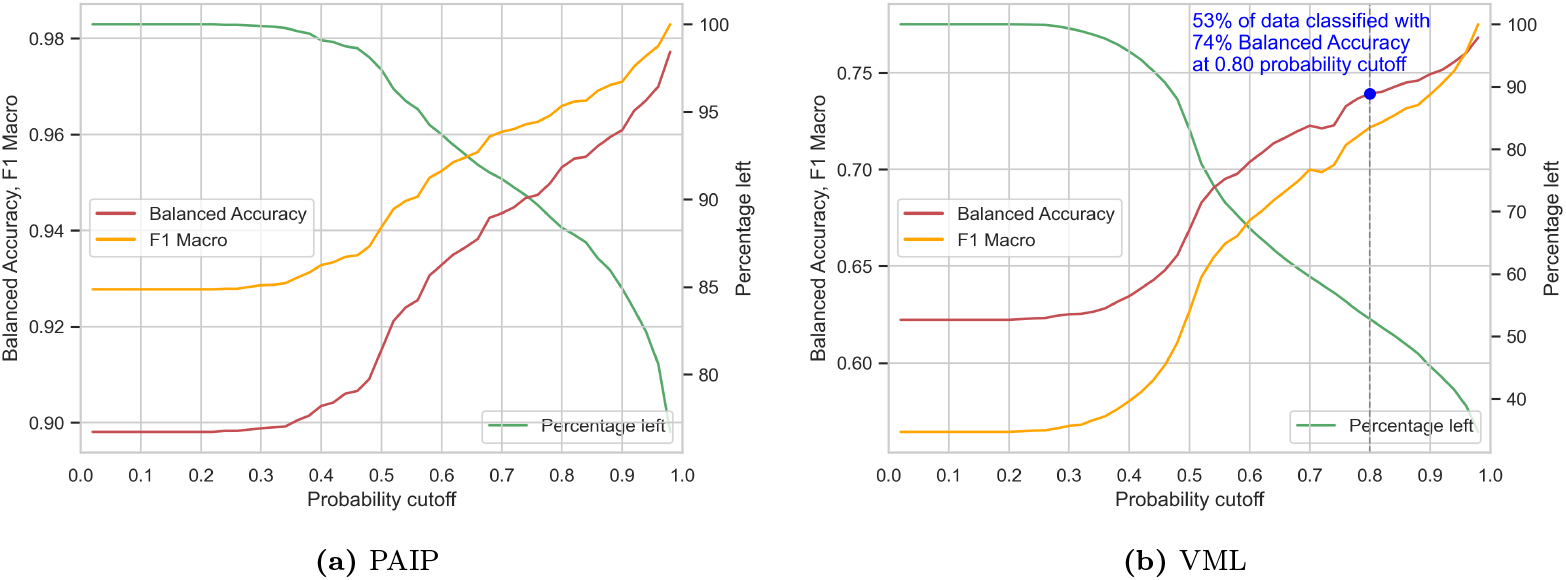
Performance in dependence of a minimum probability cutoff for PAIP (left) and VML (right). At each cutoff, only predictions where the maximum class probability is greater than or equal to the cutoff are evaluated. On the left y-axis, performance is measured via balanced accuracy and the Macro F1 metric. On the right y-axis, the corresponding percentage of the predicted data is shown.

### 3.4. Investigating wrong predictions

To assess whether high-confidence errors were due to overconfidence of the model or errors in the ground truth, we reviewed misclassifications with maximum predicted probability ≥ 0.8. For each cohort, we selected up to 10 cases with the highest predicted probability per wrongly predicted organ. A single pathologist assessed the image thumbnails as seen by the model, and if needed also the full WSI, without knowledge of the data source and any labels. For PAIP, 109 samples were high-probability misclassifications, from which we selected the top misclassifications per organ totaling 37. The pathologist agreed on 7 (19%) slides with the ground truth, agreed with the model (and not the ground truth) on 14 (38%), disagreed with both on 3 (8%), and could not determine the organ for 13 (35%), even after inspecting the full whole slide image at high magnification. For VML, 1677 samples were high probability misclassifications, from which we selected 140 for visual confirmation. The pathologist agreed with the ground truth on 52 (37%), agreed with the model and not the ground truth on 19 (14%), disagreed with both for 18 (13%), and could not determine the organ for 51 (36%) slides. Fig. 4 shows three randomly selected examples of each type.

**Figure 4.**
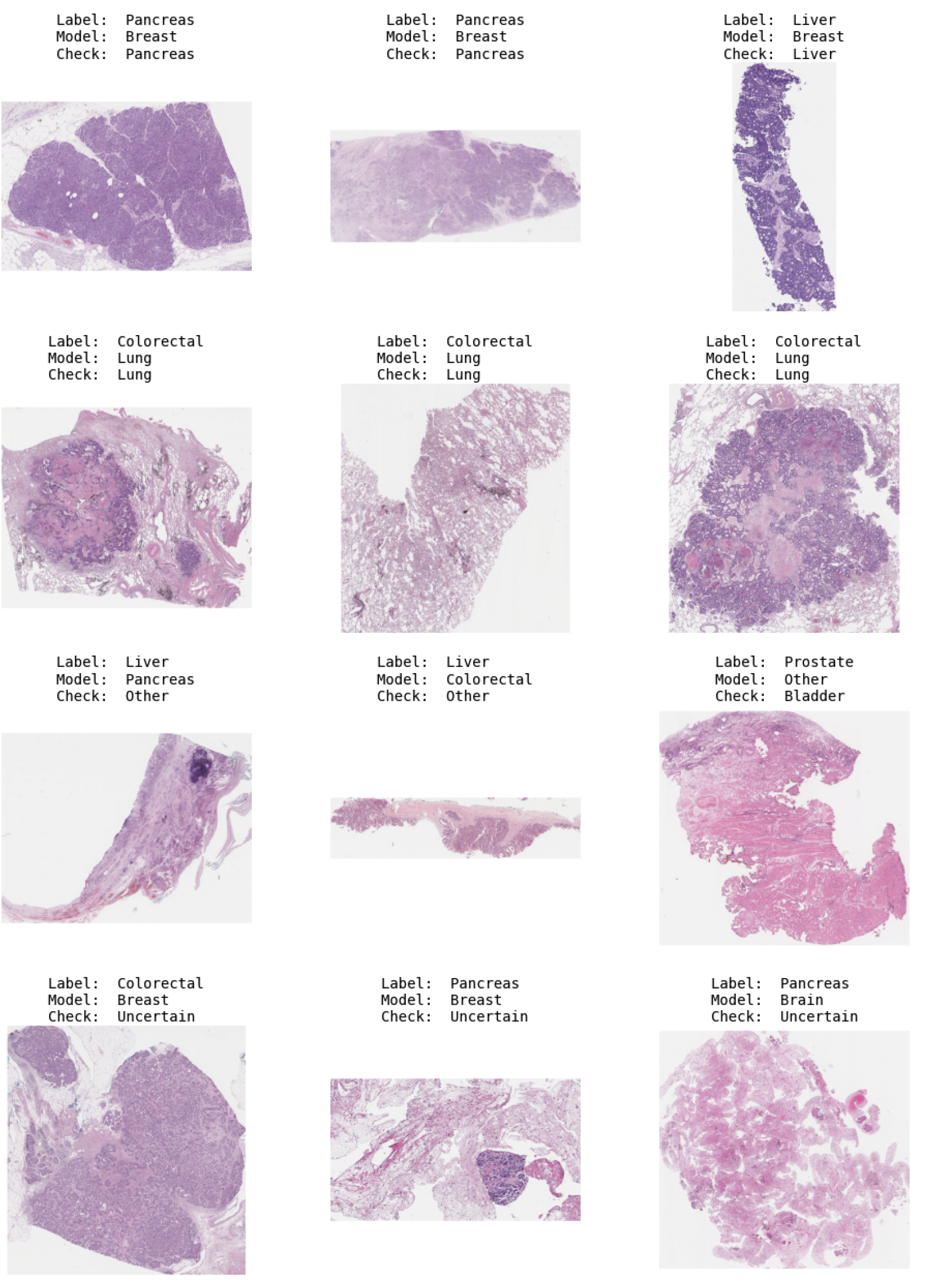
Misclassified thumbnails. Row 1: Pathologist agrees with ground truth label. Row 2: Pathologist agrees with the model. Row 3: Pathologist disagrees with both. Row 4: Pathologist could not determine the organ.

## Discussion

This work shows that organ-of-origin classification from H&E whole-slide images can be sufficiently accurate and fast to support large-scale quality control in digital pathology across a set of common human organs using only a single low-resolution thumbnail. While overall results were consistent, the model performed worse on Bladder, Skin and Stomach. Considering the different VML subsets, best performance was achieved on tumor resections. Notably, only a minor performance decrease (1.4 percentage points) was observed on non-primary resections indicating that the model can recover organ-of-origin from metastatic tissue for some cases, despite being trained mostly on primary resections. Performance on healthy resections and tumor biopsies was weak, likely because the training data included only tumor resections. In addition, biopsies often contain limited and fragmented tissue, sometimes dominated by stroma or inflammation, leaving little visible organ-specific tissue. Nevertheless, these results show that the model partly generalizes to previously unseen specimen types.

The pathologist review in Section 3.4 revealed that a portion of the model’s errors are due to a combination of errors in the ground truth (label noise), non-diagnostic tissue and genuine model failures. The disagreement between the pathologist’s conclusion and the ground truth labels indicates possible mislabeling in the evaluation data rather than pure model overconfidence. This highlights a fundamental limitation of retrospective evaluations on large public datasets, where label noise is difficult to detect and correct. Performance measurements based solely on dataset labels may therefore underestimate the true clinical usefulness of the approach. These findings suggest that organ-of-origin models could also serve as a tool for quality control even when organ information is available. Flagging high-confidence disagreements could help identify mislabeled slides or tissue contamination, or prompt manual review of ambiguous cases.

Furthermore, some slides don’t contain enough organ-specific tissue for reliable prediction of the organ of origin. This was confirmed by the visual inspection by the pathologist, where about 35% of the misclassified slides did not contain organ-specific tissue. This is more likely for small biopsies and could further explain the model’s poor performance on them.

Several limitations need to be considered. First, the current model is limited to fourteen organ classes including an *Other* category. This coarse grouping limits applicability and may affect robustness when uncommon or heterogeneous tissue types are encountered. Expanding the number of organs and including rarer entities and mixed tissues would improve the clinical applicability of the approach. Second, before full automation in a clinical setting, evaluation on independent data from multiple institutions, scanners, and staining conditions would be necessary to better assess generalizability.

At this stage, we envision the model as a quality control triage tool rather than a standalone organ verifier. Its primary use is to flag high-confidence predictions that disagree with the expected organ label for manual review before further processing, reducing review burden in large-scale digitization pipelines. The approach is therefore well suited for early integration into digital pathology workflows and is intended to complement, not replace, metadata-based checks by providing an independent visual verification step. More broadly, clinically reliable organ-of-origin classification remains a challenge. Performance is highest for common, well-preserved, diagnostically typical cases, but is limited by factors such as scant tissue and label noise in retrospective datasets. Including tissue from different magnifications and a broader range of specimen types during training would likely make the model more robust, especially on biopsies and healthy resections. Future work should also evaluate calibration and abstention more explicitly, to better identify uncertain cases for manual review. Finally, prospective validation in routine scanning workflows, ideally across multiple institutions, is needed to measure the real-world impact.

## Data Availability

The TCGA slides are available at https://portal.gdc.cancer.gov. The CPTAC slides are available at https://www.cancerimagingarchive.net. The PAIP slides are available at https://www.wisepaip. org. The VML slides are available at https://wirtualnymikroskop.mostwiedzy.pl.

## Author contributions statement

W.A. designed and conducted the experiments, wrote the software, analyzed the results and wrote the manuscript. J.W. and M.Z. assisted with data acquisition and management. Y.K. performed pathologist review of a subset of the images. M.P. contributed to software development. C.A. supervised the experiments. All authors reviewed the manuscript.

## Ethics approval and consent to participate

This study is a retrospective analysis of de-identified whole-slide images and associated metadata from publicly available datasets. No new patient data were collected and no patient contact occurred. Ethics approval and informed consent were not required for this study.

## Declaration of Generative AI and AI-assisted technologies in the manuscript preparation process

During the preparation of this work, the authors used ChatGPT 5.2 to assist with minor phrasing improvements and grammatical refinement. All content generated with this tool was reviewed, revised, and approved by the authors, who take full responsibility for the final manuscript.

