## Supplementary Material for "Fast Organ-of-Origin Classification for Digital Pathology Quality Control"

**Extended Data**

Table S1 lists the original primary site, the assigned organ class and the corresponding number of whole slide images per cohort used in this study.

| Organ | Primary Site | TCGA | CPTAC | PAIP | VML | Total |
|---|---|---|---|---|---|---|
| Bladder | Anterior wall of bladder | 0 | 0 | 0 | 1 | 1 |
| Bladder | Bladder | 458 | 0 | 0 | 114 | 572 |
| Bladder | Bladder neck | 0 | 0 | 0 | 1 | 1 |
| Bladder | Lateral wall of bladder | 0 | 0 | 0 | 1 | 1 |
| Bladder | Posterior wall of bladder | 0 | 0 | 0 | 2 | 2 |
| Bladder | Trigone of bladder | 0 | 0 | 0 | 1 | 1 |
| Brain | Brain | 1706 | 244 | 0 | 87 | 2037 |
| Brain | Brain stem | 0 | 0 | 0 | 15 | 15 |
| Brain | Cerebellum | 0 | 0 | 0 | 23 | 23 |
| Brain | Cerebrum | 0 | 0 | 0 | 229 | 229 |
| Brain | Frontal lobe | 0 | 0 | 0 | 24 | 24 |
| Brain | Parietal lobe | 0 | 0 | 0 | 15 | 15 |
| Brain | Temporal lobe | 0 | 0 | 0 | 34 | 34 |
| Breast | Axillary tail of breast | 0 | 0 | 0 | 13 | 13 |
| Breast | Breast | 1122 | 396 | 0 | 346 | 1864 |
| Breast | Central portion of breast | 0 | 0 | 0 | 183 | 183 |
| Breast | Lower-inner quadrant of breast | 0 | 0 | 0 | 102 | 102 |
| Breast | Lower-outer quadrant of breast | 0 | 0 | 0 | 86 | 86 |
| Breast | Overlapping lesion of breast | 0 | 0 | 0 | 47 | 47 |
| Breast | Upper-inner quadrant of breast | 0 | 0 | 0 | 181 | 181 |
| Breast | Upper-outer quadrant of breast | 0 | 0 | 0 | 315 | 315 |
| Colorectal | Anal canal | 0 | 0 | 0 | 2 | 2 |
| Colorectal | Anus | 0 | 0 | 0 | 13 | 13 |
| Colorectal | Appendix | 0 | 0 | 0 | 74 | 74 |
| Colorectal | Ascending colon | 0 | 0 | 0 | 53 | 53 |
| Colorectal | Cecum | 0 | 0 | 0 | 22 | 22 |
| Colorectal | Colon | 446 | 220 | 893 | 176 | 1735 |
| Colorectal | Descending colon | 0 | 0 | 0 | 25 | 25 |
| Colorectal | Hepatic flexure of colon | 0 | 0 | 0 | 5 | 5 |
| Colorectal | Overlapping lesion of colon | 0 | 0 | 0 | 7 | 7 |

*Continued on next page*

| Organ | Primary Site | TCGA | CPTAC | PAIP | VML | Total |
|---|---|---|---|---|---|---|
| Colorectal | Rectosigmoid junction | 75 | 0 | 0 | 25 | 100 |
| Colorectal | Rectum | 81 | 3 | 0 | 61 | 145 |
| Colorectal | Sigmoid colon | 0 | 0 | 0 | 50 | 50 |
| Colorectal | Transverse colon | 0 | 0 | 0 | 16 | 16 |
| Kidney | Kidney | 944 | 777 | 400 | 544 | 2665 |
| Kidney | Renal pelvis | 0 | 0 | 0 | 2 | 2 |
| Liver | Biliary | 0 | 0 | 48 | 0 | 48 |
| Liver | Liver | 0 | 0 | 556 | 546 | 1102 |
| Liver | Liver and intrahepatic bile ducts | 408 | 0 | 0 | 0 | 408 |
| Lung | Bronchus and lung | 1044 | 1526 | 0 | 0 | 2570 |
| Lung | Lower lobe, lung | 0 | 0 | 0 | 50 | 50 |
| Lung | Lung | 0 | 0 | 0 | 416 | 416 |
| Lung | Main bronchus | 0 | 0 | 0 | 1 | 1 |
| Lung | Middle lobe, lung | 0 | 0 | 0 | 12 | 12 |
| Lung | Overlapping lesion of lung | 0 | 0 | 0 | 1 | 1 |
| Lung | Trachea | 0 | 0 | 0 | 27 | 27 |
| Lung | Upper lobe, lung | 0 | 0 | 0 | 76 | 76 |
| Other | Abdomen | 0 | 0 | 0 | 11 | 11 |
| Other | Abdominal esophagus | 0 | 0 | 0 | 1 | 1 |
| Other | Accessory sinus | 0 | 0 | 0 | 16 | 16 |
| Other | Adrenal gland | 390 | 0 | 0 | 76 | 466 |
| Other | Ampulla of Vater | 0 | 0 | 0 | 2 | 2 |
| Other | Anterior 2/3 of tongue | 0 | 0 | 0 | 1 | 1 |
| Other | Anterior mediastinum | 0 | 0 | 0 | 1 | 1 |
| Other | Autonomic nervous system | 0 | 0 | 0 | 10 | 10 |
| Other | Base of tongue | 20 | 0 | 0 | 7 | 27 |
| Other | Biliary tract | 0 | 0 | 0 | 1 | 1 |
| Other | Bone | 0 | 0 | 0 | 9 | 9 |
| Other | Bone marrow | 0 | 0 | 0 | 61 | 61 |
| Other | Bone of limb | 0 | 0 | 0 | 2 | 2 |
| Other | Bones of skull and face and associated joints | 0 | 0 | 0 | 26 | 26 |
| Other | Bones, joints and articular cartilage of limbs | 4 | 0 | 0 | 0 | 4 |
| Other | Bones, joints and articular cartilage of other and unspecified sites | 2 | 0 | 0 | 0 | 2 |
| Other | Cerebral meninges | 0 | 0 | 0 | 7 | 7 |
| Other | Cheek mucosa | 0 | 0 | 0 | 1 | 1 |
| Other | Ciliary body | 0 | 0 | 0 | 18 | 18 |
| Other | Conjunctiva | 0 | 0 | 0 | 4 | 4 |
| Other | Connective, subcutaneous and other soft tissues | 431 | 0 | 0 | 22 | 453 |
| Other | Connective, subcutaneous and other soft tissues of abdomen | 0 | 0 | 0 | 69 | 69 |
| Other | Connective, subcutaneous and other soft tissues of head, face, and neck | 0 | 0 | 0 | 53 | 53 |

*Continued on next page*

| Organ | Primary Site | TCGA | CPTAC | PAIP | VML | Total |
|---|---|---|---|---|---|---|
| Other | Connective, subcutaneous and other soft tissues of lower limb and hip | 0 | 0 | 0 | 48 | 48 |
| Other | Connective, subcutaneous and other soft tissues of pelvis | 0 | 0 | 0 | 19 | 19 |
| Other | Connective, subcutaneous and other soft tissues of thorax | 0 | 0 | 0 | 101 | 101 |
| Other | Connective, subcutaneous and other soft tissues of trunk | 0 | 0 | 0 | 17 | 17 |
| Other | Connective, subcutaneous and other soft tissues of upper limb and shoulder | 0 | 0 | 0 | 48 | 48 |
| Other | Cranial nerve | 0 | 0 | 0 | 3 | 3 |
| Other | Descended testis | 0 | 0 | 0 | 26 | 26 |
| Other | Dorsal surface of tongue | 0 | 0 | 0 | 5 | 5 |
| Other | Duodenum | 0 | 0 | 0 | 27 | 27 |
| Other | Epididymis | 0 | 0 | 0 | 11 | 11 |
| Other | Esophagus | 156 | 0 | 0 | 15 | 171 |
| Other | Ethmoid sinus | 0 | 0 | 0 | 2 | 2 |
| Other | External ear | 0 | 0 | 0 | 36 | 36 |
| Other | External lower lip | 0 | 0 | 0 | 1 | 1 |
| Other | External upper lip | 0 | 0 | 0 | 1 | 1 |
| Other | Extrahepatic bile duct | 0 | 0 | 0 | 3 | 3 |
| Other | Eye and adnexa | 53 | 0 | 0 | 0 | 53 |
| Other | Eyelid | 0 | 0 | 0 | 18 | 18 |
| Other | Fallopian tube | 0 | 0 | 0 | 100 | 100 |
| Other | Female genital tract | 0 | 0 | 0 | 4 | 4 |
| Other | Floor of mouth | 57 | 0 | 0 | 14 | 71 |
| Other | Frontal sinus | 0 | 0 | 0 | 4 | 4 |
| Other | Gallbladder | 1 | 0 | 0 | 66 | 67 |
| Other | Gastrointestinal tract | 0 | 0 | 0 | 1 | 1 |
| Other | Glans penis | 0 | 0 | 0 | 4 | 4 |
| Other | Glottis | 0 | 0 | 0 | 11 | 11 |
| Other | Gum | 11 | 0 | 0 | 4 | 15 |
| Other | Hard palate | 0 | 0 | 0 | 1 | 1 |
| Other | Head, face or neck | 0 | 0 | 0 | 75 | 75 |
| Other | Heart | 0 | 0 | 0 | 205 | 205 |
| Other | Heart, mediastinum, and pleura | 123 | 0 | 0 | 0 | 123 |
| Other | Hypopharynx | 8 | 0 | 0 | 1 | 9 |
| Other | Ileum | 0 | 0 | 0 | 22 | 22 |
| Other | Intestinal tract | 0 | 0 | 0 | 3 | 3 |
| Other | Intra-abdominal lymph nodes | 0 | 0 | 0 | 207 | 207 |
| Other | Intrathoracic lymph nodes | 0 | 0 | 0 | 31 | 31 |
| Other | Jejunum | 0 | 0 | 0 | 23 | 23 |
| Other | Labium majus | 0 | 0 | 0 | 30 | 30 |
| Other | Labium minus | 0 | 0 | 0 | 8 | 8 |
| Other | Lacrimal gland | 0 | 0 | 0 | 1 | 1 |
| Other | Larynx | 111 | 0 | 0 | 124 | 235 |
| Other | Lip | 5 | 0 | 0 | 1 | 6 |

*Continued on next page*

| Organ | Primary Site | TCGA | CPTAC | PAIP | VML | Total |
|---|---|---|---|---|---|---|
| Other | Long bones of lower limb and associated joints | 0 | 0 | 0 | 18 | 18 |
| Other | Long bones of upper limb, scapula and associated joints | 0 | 0 | 0 | 1 | 1 |
| Other | Lower gum | 0 | 0 | 0 | 1 | 1 |
| Other | Lower limb | 0 | 0 | 0 | 26 | 26 |
| Other | Lymph node | 0 | 0 | 0 | 74 | 74 |
| Other | Lymph nodes | 26 | 0 | 0 | 0 | 26 |
| Other | Lymph nodes of axilla or arm | 0 | 0 | 0 | 141 | 141 |
| Other | Lymph nodes of head, face and neck | 0 | 0 | 0 | 348 | 348 |
| Other | Lymph nodes of inguinal region or leg | 0 | 0 | 0 | 53 | 53 |
| Other | Major salivary gland | 0 | 0 | 0 | 3 | 3 |
| Other | Mandible | 0 | 0 | 0 | 31 | 31 |
| Other | Maxillary sinus | 0 | 0 | 0 | 41 | 41 |
| Other | Mediastinum | 0 | 0 | 0 | 3 | 3 |
| Other | Medulla of adrenal gland | 0 | 0 | 0 | 47 | 47 |
| Other | Meninges | 1 | 0 | 0 | 0 | 1 |
| Other | Middle ear | 0 | 0 | 0 | 19 | 19 |
| Other | Mouth | 0 | 0 | 0 | 19 | 19 |
| Other | Nasal cavity | 0 | 0 | 0 | 61 | 61 |
| Other | Nasopharynx | 0 | 0 | 0 | 17 | 17 |
| Other | Nervous system | 0 | 0 | 0 | 6 | 6 |
| Other | Nipple | 0 | 0 | 0 | 28 | 28 |
| Other | Occipital lobe | 0 | 0 | 0 | 4 | 4 |
| Other | Orbit | 0 | 0 | 0 | 13 | 13 |
| Other | Oropharynx | 7 | 0 | 0 | 6 | 13 |
| Other | Other and ill-defined sites | 4 | 382 | 0 | 0 | 386 |
| Other | Other and ill-defined sites in lip, oral cavity and pharynx | 57 | 0 | 0 | 0 | 57 |
| Other | Other and unspecified major salivary glands | 1 | 0 | 0 | 0 | 1 |
| Other | Other and unspecified male genital organs | 2 | 0 | 0 | 0 | 2 |
| Other | Other and unspecified parts of biliary tract | 2 | 0 | 0 | 0 | 2 |
| Other | Other and unspecified parts of mouth | 41 | 0 | 0 | 0 | 41 |
| Other | Other and unspecified parts of tongue | 121 | 0 | 0 | 0 | 121 |
| Other | Other endocrine glands and related structures | 1 | 0 | 0 | 0 | 1 |
| Other | Other specified parts of female genital organs | 0 | 0 | 0 | 6 | 6 |
| Other | Other specified parts of male genital organs | 0 | 0 | 0 | 11 | 11 |
| Other | Other specified parts of pancreas | 0 | 0 | 0 | 2 | 2 |
| Other | Ovary | 108 | 129 | 0 | 573 | 810 |
| Other | Overlapping lesion of bones, joints and articular cartilage | 0 | 0 | 0 | 2 | 2 |
| Other | Overlapping lesion of digestive system | 0 | 0 | 0 | 2 | 2 |

*Continued on next page*

| Organ | Primary Site | TCGA | CPTAC | PAIP | VML | Total |
|---|---|---|---|---|---|---|
| Other | Overlapping lesion of heart, mediastinum and pleura | 0 | 0 | 0 | 5 | 5 |
| Other | Overlapping lesion of larynx | 0 | 0 | 0 | 3 | 3 |
| Other | Overlapping lesion of smallintestine | 0 | 0 | 0 | 10 | 10 |
| Other | Palate | 6 | 0 | 0 | 6 | 12 |
| Other | Parathyroid gland | 0 | 0 | 0 | 14 | 14 |
| Other | Parotid gland | 0 | 0 | 0 | 138 | 138 |
| Other | Pelvic bones, sacrum, coccyx and associated joints | 0 | 0 | 0 | 39 | 39 |
| Other | Pelvic lymph nodes | 0 | 0 | 0 | 197 | 197 |
| Other | Pelvis | 0 | 0 | 0 | 31 | 31 |
| Other | Peripheral nerves and autonomic nervous system | 5 | 0 | 0 | 0 | 5 |
| Other | Peritoneum | 0 | 0 | 0 | 68 | 68 |
| Other | Pharynx | 0 | 0 | 0 | 17 | 17 |
| Other | Placenta | 0 | 0 | 0 | 210 | 210 |
| Other | Pleura | 0 | 0 | 0 | 7 | 7 |
| Other | Posterior mediastinum | 0 | 0 | 0 | 34 | 34 |
| Other | Prepuce | 0 | 0 | 0 | 4 | 4 |
| Other | Pyriform sinus | 0 | 0 | 0 | 2 | 2 |
| Other | Reticuloendothelial system | 0 | 0 | 0 | 1 | 1 |
| Other | Retroperitoneum | 0 | 0 | 0 | 90 | 90 |
| Other | Retroperitoneum and peritoneum | 241 | 29 | 0 | 0 | 270 |
| Other | Rib, sternum, clavicle and associated joints | 0 | 0 | 0 | 21 | 21 |
| Other | Round ligament | 0 | 0 | 0 | 1 | 1 |
| Other | Scrotum | 0 | 0 | 0 | 5 | 5 |
| Other | Short bones of upper limb and associated joints | 0 | 0 | 0 | 1 | 1 |
| Other | Skin of lip | 0 | 0 | 0 | 7 | 7 |
| Other | Small intestine | 3 | 0 | 0 | 26 | 29 |
| Other | Soft palate | 0 | 0 | 0 | 6 | 6 |
| Other | Specified parts of peritoneum | 0 | 0 | 0 | 53 | 53 |
| Other | Spermatic cord | 0 | 0 | 0 | 24 | 24 |
| Other | Sphenoid sinus | 0 | 0 | 0 | 2 | 2 |
| Other | Spinal cord | 0 | 0 | 0 | 1 | 1 |
| Other | Spinal cord, cranial nerves, and other parts of central nervous system | 3 | 0 | 0 | 0 | 3 |
| Other | Spleen | 0 | 0 | 0 | 52 | 52 |
| Other | Subglottis | 0 | 0 | 0 | 3 | 3 |
| Other | Submandibular gland | 0 | 0 | 0 | 42 | 42 |
| Other | Supraglottis | 0 | 0 | 0 | 5 | 5 |
| Other | Testis | 254 | 0 | 0 | 179 | 433 |
| Other | Thoracic esophagus | 0 | 0 | 0 | 1 | 1 |
| Other | Thorax | 0 | 0 | 0 | 14 | 14 |
| Other | Thymus | 145 | 0 | 0 | 8 | 153 |
| Other | Tongue | 0 | 0 | 0 | 19 | 19 |

*Continued on next page*

| Organ | Primary Site | TCGA | CPTAC | PAIP | VML | Total |
|---|---|---|---|---|---|---|
| Other | Tonsil | 27 | 0 | 0 | 116 | 143 |
| Other | Tonsillar pillar | 0 | 0 | 0 | 1 | 1 |
| Other | Undescended testis | 0 | 0 | 0 | 3 | 3 |
| Other | Upper limb | 0 | 0 | 0 | 21 | 21 |
| Other | Upper respiratory tract | 0 | 0 | 0 | 2 | 2 |
| Other | Ureter | 0 | 0 | 0 | 18 | 18 |
| Other | Urethra | 0 | 0 | 0 | 6 | 6 |
| Other | Uterine adnexa | 0 | 0 | 0 | 1 | 1 |
| Other | Uvula | 0 | 0 | 0 | 1 | 1 |
| Other | Vagina | 0 | 0 | 0 | 25 | 25 |
| Other | Ventricle | 0 | 0 | 0 | 11 | 11 |
| Other | Vertebral column | 0 | 0 | 0 | 7 | 7 |
| Other | Vulva | 0 | 0 | 0 | 72 | 72 |
| Pancreas | Body of pancreas | 0 | 0 | 0 | 8 | 8 |
| Pancreas | Head of pancreas | 0 | 0 | 0 | 63 | 63 |
| Pancreas | Pancreas | 209 | 491 | 360 | 61 | 1121 |
| Pancreas | Tail of pancreas | 0 | 0 | 0 | 17 | 17 |
| Prostate | Prostate | 0 | 0 | 600 | 0 | 600 |
| Prostate | Prostate gland | 450 | 0 | 0 | 577 | 1027 |
| Skin | Skin | 418 | 0 | 0 | 90 | 508 |
| Skin | Skin of lower limb and hip | 0 | 0 | 0 | 114 | 114 |
| Skin | Skin of other and unspecified parts of face | 0 | 0 | 0 | 149 | 149 |
| Skin | Skin of scalp and neck | 0 | 0 | 0 | 99 | 99 |
| Skin | Skin of trunk | 0 | 0 | 0 | 296 | 296 |
| Skin | Skin of upper limb and shoulder | 0 | 0 | 0 | 73 | 73 |
| Stomach | Body of stomach | 0 | 0 | 0 | 19 | 19 |
| Stomach | Cardia | 0 | 0 | 0 | 38 | 38 |
| Stomach | Fundus of stomach | 0 | 0 | 0 | 8 | 8 |
| Stomach | Gastric antrum | 0 | 0 | 0 | 11 | 11 |
| Stomach | Greater curvature of stomach | 0 | 0 | 0 | 3 | 3 |
| Stomach | Lesser curvature of stomach | 0 | 0 | 0 | 10 | 10 |
| Stomach | Overlapping lesion of stomach | 0 | 0 | 0 | 15 | 15 |
| Stomach | Pylorus | 0 | 0 | 0 | 12 | 12 |
| Stomach | Stomach | 379 | 0 | 0 | 62 | 441 |
| Thyroid gland | Thyroid gland | 508 | 0 | 0 | 478 | 986 |
| Uterus | Broad ligament | 0 | 0 | 0 | 32 | 32 |
| Uterus | Cervix uteri | 279 | 0 | 0 | 34 | 313 |
| Uterus | Corpus uteri | 513 | 0 | 0 | 269 | 782 |
| Uterus | Endocervix | 0 | 0 | 0 | 98 | 98 |
| Uterus | Endometrium | 0 | 0 | 0 | 150 | 150 |
| Uterus | Exocervix | 0 | 0 | 0 | 453 | 453 |
| Uterus | Fundus uteri | 0 | 0 | 0 | 4 | 4 |
| Uterus | Isthmus uteri | 0 | 0 | 0 | 35 | 35 |
| Uterus | Myometrium | 0 | 0 | 0 | 77 | 77 |
| Uterus | Overlapping lesion of cervix uteri | 0 | 0 | 0 | 2 | 2 |
| Uterus | Parametrium | 0 | 0 | 0 | 36 | 36 |
| Uterus | Uterus | 112 | 848 | 0 | 26 | 986 |

*Continued on next page*

| Organ | Primary Site | TCGA | CPTAC | PAIP | VML | Total |
|---|---|---|---|---|---|---|
| Total | | 11579 | 5045 | 2857 | 12348 | 31829 |

**Table S1:** Number of whole slide images per organ and dataset.
